# Robust induction of B cell and T cell responses by a third dose of inactivated SARS-CoV-2 vaccine

**DOI:** 10.1101/2021.09.12.21263373

**Authors:** Yihao Liu, Qin Zeng, Caiguanxi Deng, Mengyuan Li, Liubing Li, Dayue Liu, Ming Liu, Xinyuan Ruan, Jie Mei, Ruohui Mo, Qian Zhou, Min Liu, Sui Peng, Ji Wang, Hui Zhang, Haipeng Xiao

## Abstract

SARS-CoV-2 inactivated vaccines have shown remarkable efficacy in clinical trials, especially in reducing severe illness and casualty. However, the waning of humoral immunity over time has raised concern over the durability of immune memory following vaccination. Thus, we conducted a non-randomized trial among the healthcare professionals (HCWs) to investigate the long-term sustainability of SARS-CoV-2-specific B cells and T cells stimulated by inactivated vaccines and the potential need for a third booster dose. Although neutralizing antibodies elicited by the standard two-dose vaccination schedule dropped from a peak of 29.3 AU/ml to 8.8 AU/ml 5 months after the second vaccination, spike-specific memory B and T cells were still detectable, forming the basis for a quick recall response. As expected, the faded humoral immune response was vigorously elevated to 63.6 AU/ml by 7.2 folds 1 week after the third dose along with abundant spike-specific circulating follicular helper T cells in parallel. Meanwhile, spike-specific CD4^+^ and CD8^+^ T cells were also robustly elevated by 5.9 and 2.7 folds respectively. Robust expansion of memory pools by the third dose potentiated greater durability of protective immune responses. Another key finding in this trial was that HCWs with low serological response to 2 doses were not truly “non-responders” but fully equipped with immune memory that could be quickly recalled by a third dose even 5 months after the second vaccination. Collectively, these data provide insights into the generation of long-term immunological memory by the inactivated vaccine, which could be rapidly recalled and further boosted by a third dose.

## Introduction

The coronavirus disease 2019 (COVID-19), caused by the severe acute respiratory syndrome coronavirus 2 (SARS-CoV-2), continues to spread across the globe currently^1, 2^. The pandemic has brought profound casualties of human lives and socioeconomic issues. The establishment of herd immunity by vaccination represents the most cost-effective strategy to prevent COVID-19. The rapid spreading of COVID-19 has urged the governments to authorize the emergent use of vaccines against SARS-CoV-2^3–5^.

The presence of neutralizing antibodies (NAbs) against SARS-CoV-2 is an indicator of protective immunity after vaccination or infection^6, 7^. NAbs capable of blocking the interaction between the spike protein and its receptor angiotensin converting enzyme 2 (ACE2) are particularly important for protection from COVID-19^8^. Therefore, inducing potent NAbs and long-lasting memory B cells are the primary goal of SARS-CoV-2 vaccines. Two doses of mRNA or inactivated vaccines are capable of inducing potent neutralizing antibody responses ^3, 9, 10^. Our previous study further demonstrated that inactivated vaccines elicited SARS-CoV-2 specific memory B cells^11^, which are important for a rapid and robust recall of protective responses against viral infection. However, little is known how long can these immune responses sustain. A rapid decline of neutralizing antibodies has been observed among infected healthcare workers (HCWs)^12^. Neutralizing antibodies also waned overtime after the second dose of BNT162b2 or ChAdOx1^13^, indicating weakened protection for SARS-CoV-2 infection. In addition, the rapid emergence of novel SARS-CoV-2 variants of concern (VOCs) dampens the efficacy of SARS-CoV-2 vaccines, since vaccine-induced antibodies were less effective in neutralizing some emerging VOCs^14^. Moreover, cross-reactive antibodies against VOCs decayed even faster than antibodies against wild-type strain^15^.

Besides humoral immune responses, T cells also play a pivotal role in coordinating the adaptive immune responses and as effectors against viral infection. In some cases, patients with inherited or treatment-induced B cell deficiency were able to recover from COVID-19^16, 17^, suggesting a potential role of cellular responses in fighting against SARS-CoV-2 infection. Induction of CD8^+^ T cell responses was documented in both SARS-CoV-2 infection and vaccination^18, 19^. On the other hand, rapid induction of CD4^+^ T cells is associated with coordinated humoral and cellular response to a SARS-CoV-2 mRNA vaccine^20^. SARS-CoV-2 specific circulating T follicular helper cells (cTFH) cells, whose counterparts in lymph nodes reinforce B cells and humoral responses^21^, correlated with neutralizing antibody levels in convalescent COVID-19 patients^22, 23^.

Nevertheless, T cell responses induced by inactivated vaccines are much less well characterized than humoral immunity. One previous study revealed that the inactivated vaccine could elicit T cells response after 2 doses of vaccination^24^, but the phenotype and sustainability of antigen-specific CD4^+^ and CD8^+^ T cells remained unclear yet.

In this study, we sought to investigate the sustainability of immune memory stimulated by the inactivated vaccine and profile humoral and cellular responses to a third dose among HCWs. Although NAbs declined substantially, SARS-CoV-2-specific memory B, CD4^+^, and CD8^+^ T cells persisted in the peripheral blood 5 months after the second vaccination, even in participants who were seronegative after receiving 2 doses of the inactivated vaccines. The third vaccination robustly recalled both humoral and cellular immune responses in all participants.

## Materials and methods

### Human subjects

In this study, we conducted a non-randomized trial and recruited participants from a prospective cohort at the First Affiliated Hospital of Sun Yat-sen University (FAH-SYSU) in Guangzhou, China. As we described previously^11^, 63 HCWs received the inactivated SARS-CoV-2 vaccine BBIBP-CorV (BBIBP-CorV, Sinopharm, Beijing) in the morning (9 am – 11 am) or afternoon (3 pm – 5 pm) on day (d) 0 and d28, respectively. Fifty of the 63 HCWs were volunteered to receive a third booster shot of the inactivated vaccine 6 months after the prime vaccination (d180). They were assigned to the morning or afternoon group as to their previous vaccinating time accordingly. Demographic characteristics of the HCWs were summarized in Supplementary Table 1. Blood samples were collected on d180 before the booster dose and d187, d194 and d208 after the booster dose. Convalescent patients who had recovered from SARS-CoV-2 infection were recruited as the positive control (Supplementary Table 2). All studies were approved by the Institutional Review Board of FAH-SYSU and written consent was obtained from all participants. The prospective cohort and the trial were registered to Chinese Clinical Trial Registry (ChiCTR2100042222 and ChiCTR2100048665).

### Cell isolation

Blood samples were collected into the heparinized tubes and processed right after sample collection. Peripheral blood mononuclear cells (PBMCs) were isolated using density-gradient centrifugation. Briefly, blood samples were diluted with PBS at 1:1 ratio and loaded on top of Lymphoprep™ (StemCell) in the Falcon tubes. The falcon tubes were then centrifuged at 1500 rpm for 30 mins. The medium layer was collected and washed with PBS twice. PBMCs were cryopreserved in Bambanker (StemCell) immediately.

### SARS-CoV-2 neutralizing antibodies measurement

A one-step competitive Chemiluminescent immunoassay was used to detect the concentration of NAbs against SARS-CoV-2 in sera by iFlash 2019-nCoV NAb kit (YHLO Biotech Co, Ltd) as previously reported^25^. In this assay, receptor binding domain (RBD) of the SARS-CoV-2 was coated on magnetic beads. Acridinium ester-labeled ACE2 was designed to compete with SARS-CoV-2 NAbs in sera for the RBD. NAb titers were calculated by an iFlash3000 Chemiluminescence Immunoassay Analyzer (YHLO Biotech Co, Ltd). Neutralizing activity is determined in arbitrary units (AU) and the cut-off is 10 AU/ml.

### Ex vivo ELISpot assay

IFNγ ELISpot assays were performed as described previously^26^. Briefly, 100 μl of the coating antibody (15 μg/ml, MabTech, #3420-3-250) was added to the well of ELISpot plate (MabTech) and incubated overnight at 4°C. The peptide pool of SARS-CoV-2 spike protein (MabTech, #3630-1) was added to 300,000 PBMCs per test at a final concentration of 2 μg/ml for 24 h. Unstimulated cells were used as negative control while anti-CD3/CD28 dynabeads (Thermo Fisher) stimulated cells were set as the positive control. Plates were incubated with IFNγ detection antibody (1μg/ml, MabTech, #3420-6-250), followed by Avidin-HRP (1μg/ml, Vector, #A-2004-5) and visualized using the ACE substrate. Antigen-specific T cell responses were quantified by subtracting the number of spots in unstimulated cells from the peptide stimulated cells.

SARS-CoV-2-specific B cell ELISpot was performed as previously described^27^. Briefly, ELISpot plates were coated with 10 ug/ml recombinant RBD protein (Sino Biological, #40592-VNAH-100) and 8 ug/ml spike protein (Sino Biological, #40589-V08B1) overnight at 4°C. To optimize the human IgG B cell ELISpot assay, PBMCs were cultured with a mixture of R848 (1 μg/ml, Tocris, #4536/10) and IL-2 (20 ng/ml, PeproTech, #200-02-10) for 3 days to secrete a detectable amount of antibody^28, 29^. After pre-stimulation, cells were washed extensively to remove secreted antibodies. 500,000 PBMCs were added to the coated plates and incubated for 18 h at 37°C with 5% CO_2_. The following day, cells were removed and plates were incubated with Biotin-anti-Human IgG (1μg/ml, Jackson ImmunoResearch, #709-065-098). Plates were then incubated with Avidin-HRP followed by visualization with ACE substrate. ELISpot plates were analyzed using an ELISpot counter (Cellular Technologies Ltd). Results were expressed as spot-forming units (s.f.u.) per 10^6^ PBMCs. Responses were considered positive if the results were at least three times the mean of the negative control wells and >25 s.f.u. per 10^6^ PBMCs. Data were excluded when negative control wells had >30 s.f.u. per 10^6^ PBMCs or positive control wells (anti-CD3/CD28 dynabeads) had no spot.

### Activation-induced markers (AIM) T cell assay

PBMCs were cultured in RPMI 1640 supplemented with 10% FBS and 1% penicillin and streptomycin (Thermo Fisher) at 37 °C overnight. The cells were then cultured with or without the peptide pool of SARS-CoV-2 spike protein (2 μg/ml) for 12 h. An equal concentration of DMSO in PBS was used as the negative control. Antigen-specific T cells were measured as a percentage of AIM^+^ (OX40^+^CD137^+^) for CD4^+^ T or (CD69^+^CD137^+^) for CD8^+^ T cells after stimulation of PBMCs with the peptide pool of spike protein^26^. After the stimulation, cells were first stained with Zombie Red for dead cell exclusion. Cells were then stained with anti-CD3-Pacific blue, anti-CD4-BV510, anti-CD8-Percp, anti-CD69-Super Bright 436, anti-CD134 (OX40)-BV605 and anti-CD137-(4-1BB)-PE antibodies for antigen-specific T cell analysis. As for phenotype analysis of SARS-CoV-2-specific T cells, cells were further labeled with anti-CCR7-APC/cy7, anti-CD45RA-BV650, anti-CXCR5-BV711 and anti-PD-1-PE/cy7 antibodies. All FACS antibodies were from Biolegend. Stimulation with anti-CD3/CD28 dynabeads was included as positive controls. Any sample with a low response to anti-CD3/CD28 stimulation was excluded as quality control for the samples. Data were acquired by flow cytometry.

### Detection of SARS-CoV-2-specific memory B cells

SARS-CoV-2-specific memory B cells were detected as we described previously^11^. First, biotinylated antigens were multimerized with fluorescence-labeled streptavidin individually. Recombinant spike protein (R&D, #BT10549-050) was mixed with BV510-streptavidin (BioLegend) at 10:1 ratio and BV785-streptavidin (BioLegend) at 18:1 ratio at 4°C for 1h. Recombinant RBD protein (R&D, #BT10500-050) was mixed with BV421-streptavidin (BioLegend) at 20:1 ratio at 4°C for 1h. The antigen probes prepared above were then mixed in 50mM free d-biotin (Macklin) in PBS to ensure minimal cross-reactivity. PBMCs were thawed and let to rest at 37°C with 5% CO2 for 2h and stained with Zombie Red (BioLegend) in PBS at room temperature for 20 min. Cells were then stained with 50 μl of antigen probe cocktail containing 100 ng of spike and 50 ng of RBD at 4°C for 30 min. Cells were washed with PBS and then stained with the following antibody cocktail: anti-CD3-Pacific Blue™, anti-CD19-PE-CY7, anti-CD27-AF700, anti-IgD-FITC, anti-CD38-BV650, anti-IgM-BV605, anti-IgG-AF647, anti-IgA-PE all from BioLegend at 1:100 dilution. Samples were acquired by flow cytometry.

### Detection of SARS-CoV-2-specific IFNγ-producing T cells

The detection of spike-reactive T cells was performed as described with modification^30^. Briefly, overnight-rested PBMCs were cultured with or without the peptide pool of SARS-CoV-2 spike protein (4 μg/ml) in the presence of 3 μg/ml of anti-CD28 monoclonal antibodies for 24 h. Cells were then incubated with Brefeldin A for additional 5 h. Cells were stained with anti-CD3-Pacific blue, anti-CD4-BV510, anti-CD8-Percp antibodies for surface markers. Cells were then washed, fixed with Cytofix/Cytoperm and stained with anti-IFNγ-PE-Cy7 (BioLegend). Dead cells were excluded by Zombie Red staining. Samples were analyzed by flow cytometry. Samples without peptide stimulation were used as the negative control.

### Flow cytometry

All flow cytometry samples were analyzed using cryopreserved cells which were thawed and suspended in RPMI 1640 media supplemented with 2% FBS. All samples were analyzed by flow cytometry with Cytek^TM^ AURORA. FlowJo (Tree Star, USA) software was used for FACS data analysis. Details of antibodies used in this study are listed in Supplementary Table 3.

### ELISA-based antibody-antigen distribution analysis

All SARS-CoV-2 proteins were purchased from Sino Biological (Beijing, China). 200ng/well of SARS-CoV-2 spike (40589-V08B1), spike S1 subunit (40591-V08H), spike NTD (40591-V49H), spike S2 subunit (40590-V08B), RBD (40592-V08H), RBD T478K (40592-V08H91), nucleocapsid (40588-V08B) and envelope (40609-V10E3) were coated on the 96-well ELISA plate overnight at 4 °C, respectively. Plates were washed by PBS supplemented with 0.05% Tween-20 (PBST) three times, followed by blocking with 5% BSA in PBST for 1 h at room temperature. Sera were diluted 20-fold for the first well and 4-fold serial diluted for subsequent wells in 5% BSA in PBST, and incubated at 4°C overnight. Plates were washed 3 times by PBST, and added with HRP-conjugated goat anti-human IgG antibody (2040-05, SouthernBiotech, 1:3000) for 30 min at room temperature. Plates were washed five times with PBST. 3,3′,5,5′-Tetramethylbenzidine (TMB) substrate (P0209, Beyotime) was added for 15 min, and stopped by the stopping buffer (C1058, Solarbio). OD450 was measured by Varioskan Lux Microplate Reader (Thermo Fisher).

### Pseudovirus neutralizing assay

For the generation of SARS-CoV-2-Spike (Wuhan-Hu-1) pseudovirus ^31^, pcDNA3.1-2019-nCoV-Spike and pNL4-3 R⁻E⁻ (gifts from Dr. Lu Lu, Fudan University) were co-transfected to HEK293T with Lipo8000 (C0533, Beyotime) according to the manufacturer’s instruction. For the generation of the B.1.617.2 Delta-variant spike pseudovirus, pCMV3-SARS-CoV-2-Spike (VG40804-UT, Sino Biological), pSPAX2 and pLenti-CMV-Luc-puro were co-transfected to HEK293T. The virus was harvested 72 hours post-transfection and stored at −80°C until use. 2×10^4^ hACE2-293T/well were seeded on the black flat-bottom 96-well plate (655090, Greiner Bio-one) for 16 hours in advance. Sera were diluted 10-fold for the first well and 4-fold serial diluted for subsequent wells in DMEM, then co-incubated with pseudovirus at 37°C for 1 hour. Then the co-incubated samples, together with samples without sera or pseudovirus as controls, were subjected with 10μg/ml polybrene (C0351, Beyotime) to the hACE2-293T for 6-hour absorption, following by replacement of the culture medium for the next 42 hours incubation at 37°C, 5% CO₂. Infected cells were processed to the luciferase assay using the Luciferase Assay System (E4550, Promega). The illuminescence was measured by Varioskan Lux Microplate Reader (Thermo Fisher). The 50% pseudovirus neutralization titer (pNT50) were determined by four-parameter nonlinear regression curve (GraphPad Prism).

### Statistical analysis

Statistical analysis was performed using Prism 5.0. Comparisons were assessed using Wilcoxon rank sum test, Student’s t-test, paired Student’s t-test or One-way ANOVA followed by Bonferroni’s multiple comparison post-test as appropriate. P values < 0.05 were considered as statistically significant.

## Results

### Neutralizing antibody response is enhanced by a third dose of inactivated SARS-CoV-2 vaccine BBIBP-CorV

We have previously conducted a non-randomized trial and recruited HCWs from a prospective cohort, demonstrating the impact of the circadian rhythm on the immune response induced by a primary 2-dose series of the inactivated SARS-CoV-2 vaccine (BBIBP-CorV, Sinopharm, Beijing)^11^. Fifty HCWs from this cohort were volunteered to participate in the current non-randomized trial to investigate the duration of the primary vaccination regimen and potential benefits of a third dose, given 5 months after the second dose (Fig. 1A). No severe side effects related to vaccination were recorded during the trial (Supplementary Table 1).

**Figure 1.**
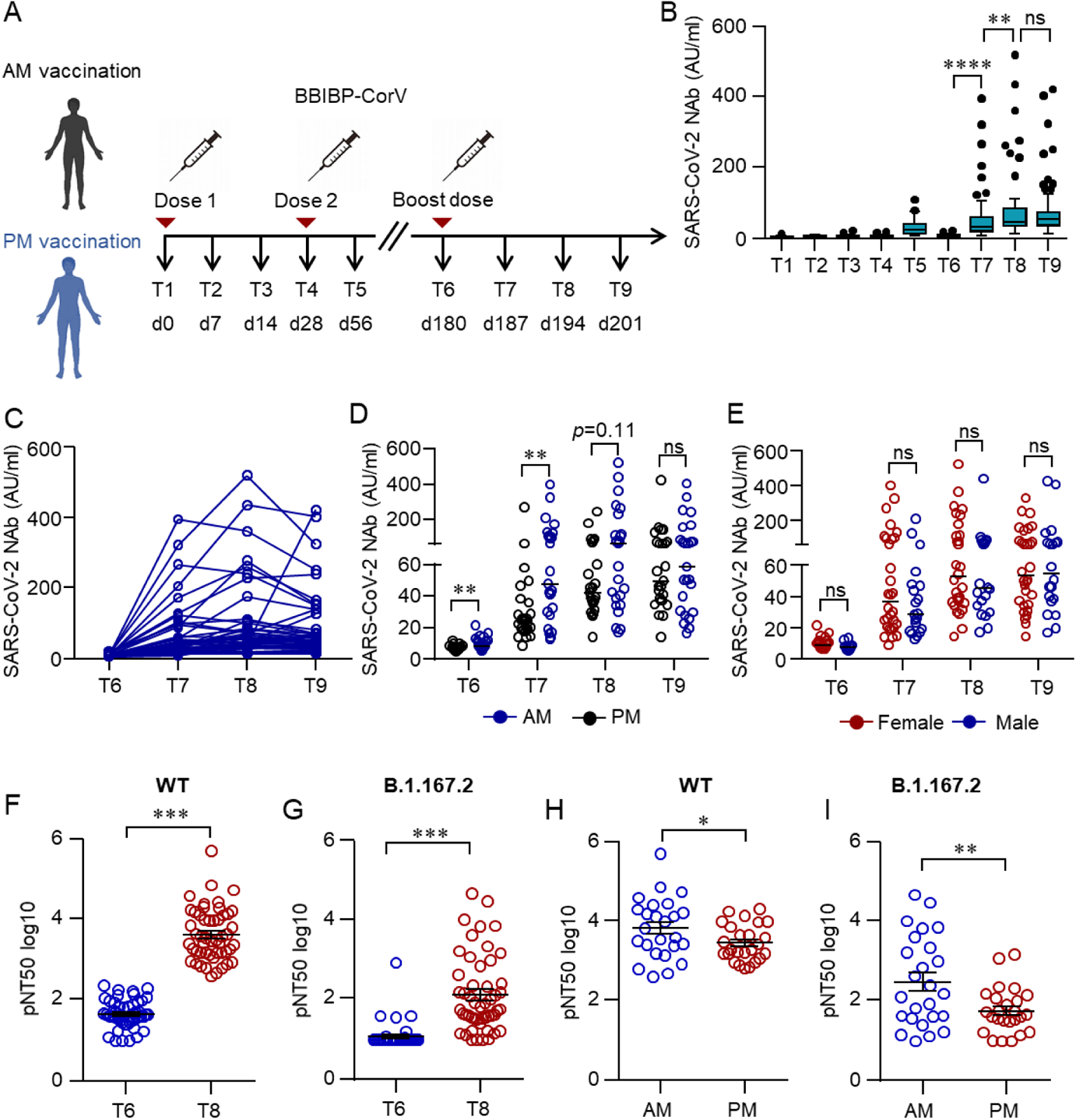
Humoral responses induced by a third dose of inactivated SARS-CoV-2 vaccine BBIBP-CorV. (A) Protocol of the prospective cohort and the non-randomized trial. Healthcare workers (HCWs) were recruited from a perspective cohort who received two doses of an inactivated vaccine either in the morning (AM, n=25) or afternoon (PM, n=25) on day (d) 0 and d28. They were assigned to morning or afternoon vaccination to receive a third dose of the inactivated vaccine on day 180 according to their previous vaccinating time. Blood samples were collected on different time points d0 (T1), d14 (T2), d21 (T3), d28 (T4), d56 (T5), d180 (T6), d187 (T7), d194 (T8) and d208 (T9). (B-E) Neutralizing antibodies (NAbs) against SARS-CoV-2 in the serum of each volunteer was measured by Chemifluorescence Assay longitudinally. (B) The concentration of NAbs in the sera at different time points was summarized and shown in the box plot. (C) The longitudinal changes of NAbs in the sera before and after the third dose of vaccination. (D) NAbs in the sera from the morning or afternoon group at different time points. (E) NAbs concentrations in the sera of female or male. Medians of the data were shown. (F, G) Neutralizing activities of sera from T6 and T8 against SARS-CoV-2 wild type (WT) and Delta variant (B.1.167.2) were measured by pseudovirus neutralizing assay. (H, I) Comparison of neutralizing activities between the morning and afternoon group on T8. Data are mean ± SEM. Comparisons were done by Wilcoxon rank sum test in B, D, E or Student’s t-test in F-I. **p<0.01, ****p<0.0001; ns, not significant.

Blood samples were collected from multiple time points before and after each dose and NAbs targeting RBD were measured by Chemiluminescent immunoassay^25^. The mean serum concentration of NAbs dropped by 70% from d56 (1 month post the second dose) to d180, 5 months after the second dose (Fig. 1B). Intriguingly, NAbs maintained a significantly higher level in the morning vaccination group than that in the afternoon group on d180 (Fig. 1D).

Although serum NAbs in 72% of participates (36/50) dropped below the threshold (10 AU/ml) on d180, serological responses could be rapidly recalled to an unprecedented level within 1 week after the third dose. The mean concentration of NAbs was increased by 7.2 folds from 8.8 AU/ml on d180 to 63.6 AU/ml on d187, peaking at 92.3 AU/ml 2 weeks after the booster (Fig. 1C).

As the primary vaccination series^11^, the circadian rhythm also governed the outcome of the third dose, albeit only at the early stage. Significantly higher levels of NAbs were observed in the morning group after the third dose on d187 (Fig. 1D). However, the difference between the morning and the afternoon groups gradually diminished over time (Fig. 1D). Moreover, we did not observe any difference in NAbs levels between females and males (Fig. 1E).

Pseudovirus neutralizing assays were next applied to confirm the elevation of serum NAbs against wild type (WT) SARS-CoV-2 and VOCs by the third dose. Consistent with the results of Chemiluminescent immunoassay, the 50% pseudovirus neutralization titer (pNT50) against WT was greatly boosted (Fig. 1F). Encouragingly, the third dose not only elevated the NAbs against WT, but also strengthen a cross-protective immune response. The mean serum pNT50 against B.1.167.2 Delta variant was increased by 10-fold from day180 to day194 (Fig. 1G). In pseudovirus assay, a significantly higher level of NAbs against both WT and B.1.167.2 could still be observed in morning vaccination participants (Fig. 1H and I).

One potential advantage of inactivated vaccines over other vaccine types is that they comprise all viral structural proteins which may induce a broader spectrum of immunity in addition to NAbs against RBD. To profile the antibody spectrum induced by BBIBP-CorV, serum IgG against major SARS-CoV-2 structural proteins and their subdomains, including S1, S2, RBD, NTD of spike protein, nucleocapsid, and small envelope protein, were measured by indirect ELISA before and after the third dose. As expected, BBIBP-CorV successfully elicited antibodies against all structural proteins and subdomains tested (Supplementary Fig. 1). Notably, we found all vaccinees were equipped with a high titer of anti-envelope IgG which has not been reported in a vaccine study elsewhere and was hardly induced even during natural SARS-CoV-2 infection^32^. Interestingly, the third dose did not elevate antibody responses against all tested structural proteins or subdomains evenly, but profoundly augment the immune response against the S1 domain of spike protein which comprises NTD and RBD subdomains (Supplementary Fig. 1).

### SARS-CoV-2 specific memory B cell response is robustly induced by a third dose of inactivated vaccine

The rapid and robust recall of humoral immune responses by the third vaccination indicated that the primary 2-dose vaccination regimen has established a sustained immune memory. We explored the presence of antigen-specific memory B cells before and after the third dose by flow cytometry.

Total B cell populations were first characterized by flow cytometry (Supplementary Fig. 2). Only mild changes in IgD^+^, IgM^+^, IgA^+^ or IgD^−^CD27^−^ B cells were found before and after the booster, whereas the percentage of total and naïve B cells were not affected (Supplementary Fig. 3), consistent with a good safety profile of the third dose (Supplementary Table. 1).

Next, antigen-specific B cell populations were investigated. Spike- and RBD-specific memory B cells could be detected at 4 weeks (d56) and 5 months (d180) after the second dose, revealing that memory B cells persisted despite a decreasing trend over a 5-months time frame between second and third immunization (Fig. 2A and C). The third dose increased the percentages of spike- and RBD-specific memory B cells by 1.7 and 2.0 folds respectively from d180 to d187, resulting in a significantly larger memory pool on d187 than that on d56 (Fig. 2B and D). The majority of these antigen-specific memory B cells could be classified into IgG^+^ or IgM^+^ B cells, while a minor population of them was IgA^+^ B cells (Fig. 2E and F).

**Figure 2.**
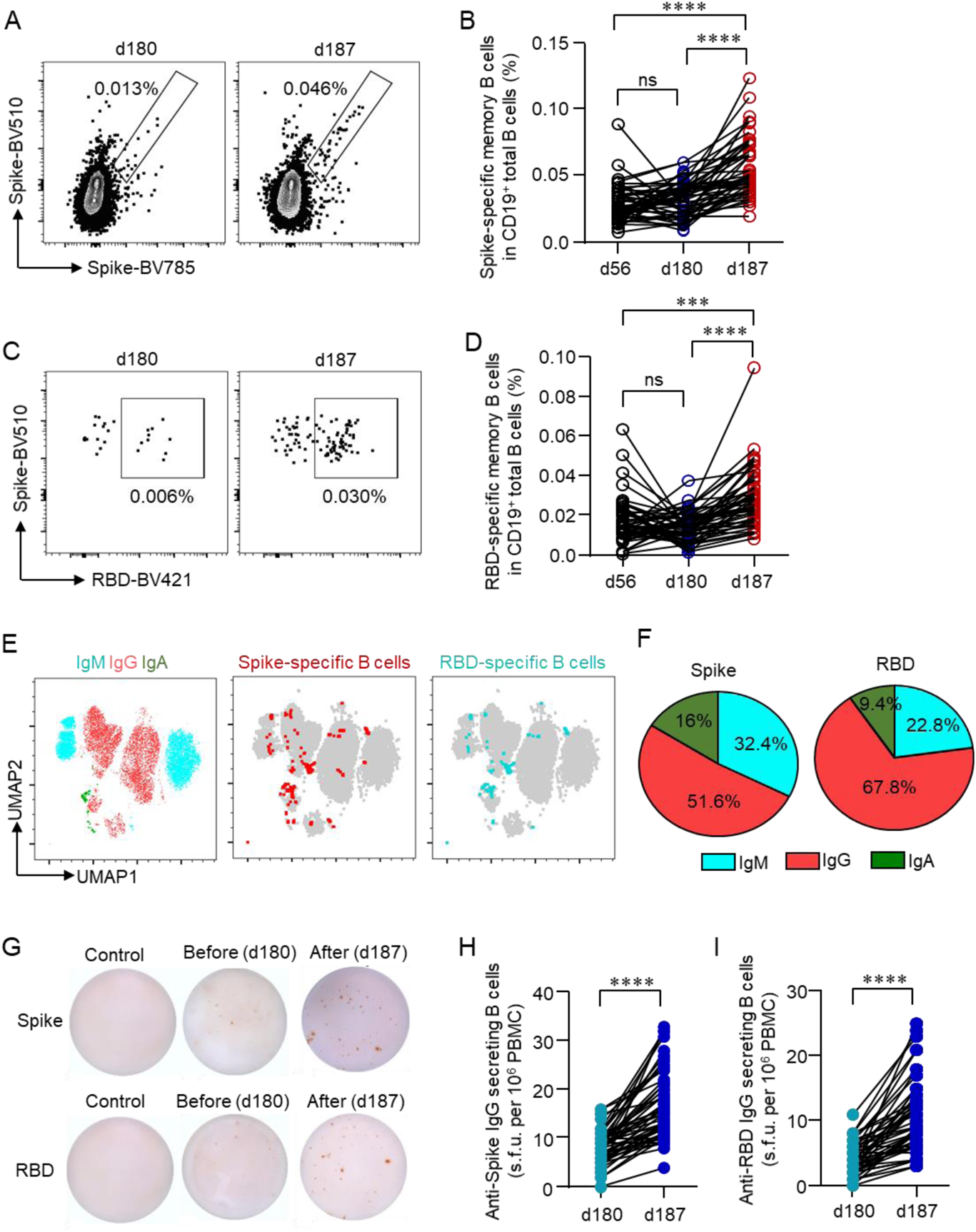
SARS-CoV-2-specific memory B cell after a third dose of inactivated vaccine. Recombinant spike or receptor binding domain (RBD) proteins were used to label SARS-CoV-2-specific BCRs. Data were acquired by flow cytometry. (A, C) Representative FACS plots for spike- or RBD-specific BCR were shown. (B, D) Longitudinal changes of spike- or RBD-specific memory B cells were summarized. (E) IgG^+^, IgA^+^ and IgM^+^ B cells were measured by flow cytometry. UMAP plot demonstrated the distribution of spike- and RBD-specific memory B cells in IgG^+^, IgA^+^ and IgM^+^ B cells. (F) Mean percentages of each BCR subtype in spike- and RBD-specific B cells were summarized. (G) Spike- or RBD-specific IgG-secreting B cells were measured by ELISpot after incubating PBMCs in spike- or RBD-coated plates for 18 h. Representative ELISpot images were shown. (H, I) Numbers of Spike- or RBD-specific IgG-secreting B cells per 10^6^ PBMCs were summarized. Comparisons were done by one-way ANOVA corrected for multiple comparison in panels B, D and Student’s paired-T test in panels H and I. ***p<0.001, ****p<0.0001; ns, not significant.

The capability of these Spike- or RBD-specific memory B cells in secreting antibodies upon the antigen stimulation was further confirmed by B cell ELISpot. The third dose boosted spike- or RBD-responsive, IgG-secreting B cells from 8 u.f.c/10^6^ PBMCs to 17 u.f.c/10^6^ PBMCs or 4 u.f.c/10^6^ PBMCs to 10.7 u.f.c/10^6^ PBMCs respectively (Fig. 2G-I). These results demonstrated that the primary 2-dose vaccination regimen had equipped vaccinees with long-lasting memory B cells which could be immediately recalled and further expanded by a third dose of the inactivated vaccine.

### The cellular immune response is boosted by a third dose of inactivated SARS-CoV-2 vaccine

Besides humoral immune responses, the cellular arm of immunity confers another layer of protection, which may particularly crucial when B cell epitopes mutate rapidly and continuously. Therefore, antigen-specific T cells induced by the primary vaccination regimen and the third dose booster were next investigated. Blood samples collected on d180 and d187 were stimulated by the spike peptide pool and IFNγ-secreting cells were quantified by ELISpot. Surprisingly, antigen-responsive IFNγ-secreting T cells were readily detected in all vaccinees even 5 months after the second dose, though a lower amount was found in some participants (Fig. 3A and B). As IgG-secreting B cells, IFNγ-secreting T cell responses were also enhanced by 2.3 folds after the third dose (Fig. 3A and B).

**Figure 3.**
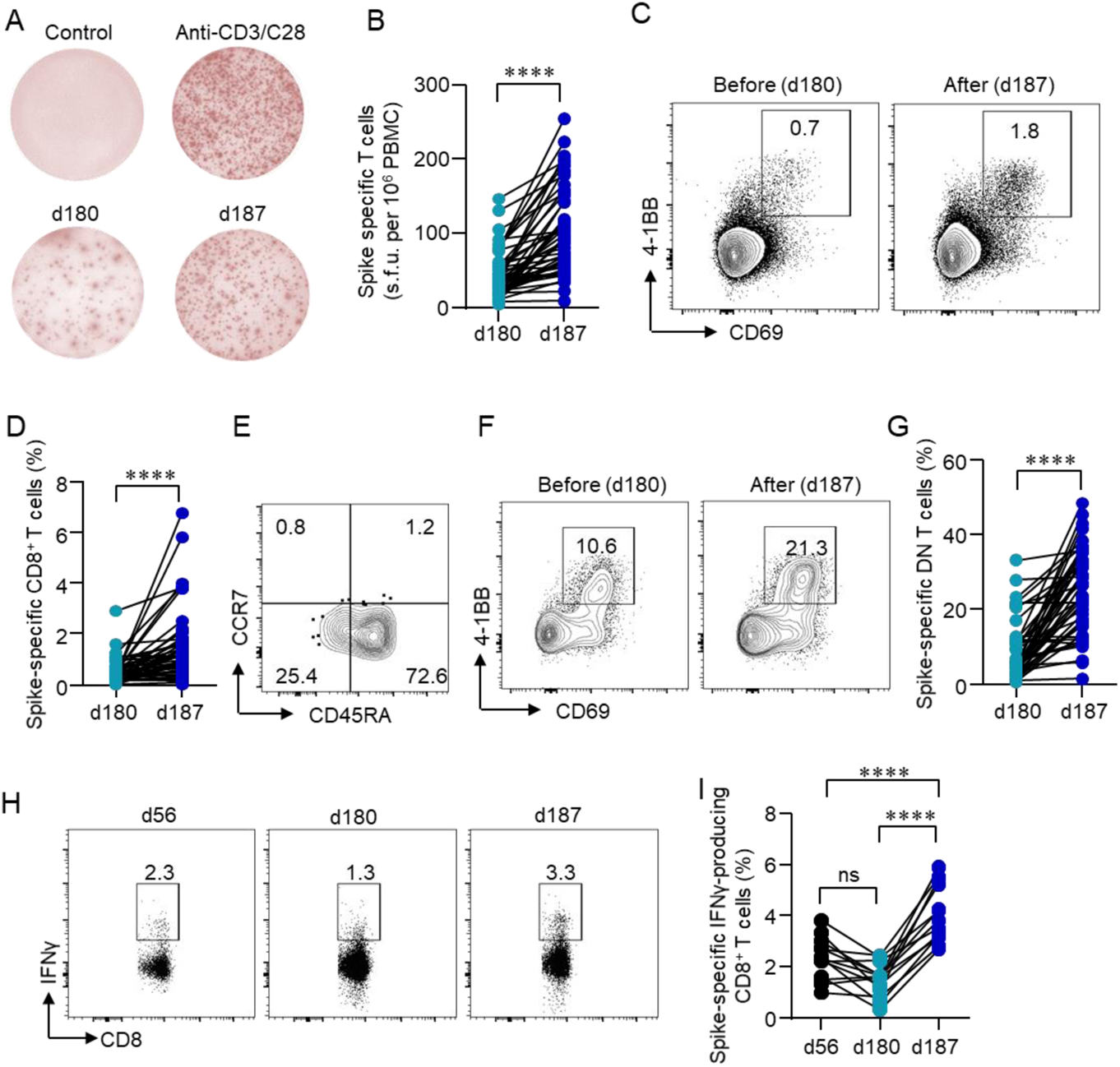
Cellular responses induced by a third dose of inactivated SARS-CoV-2 vaccine. (A) PBMCs were stimulated with or without the spike peptide pool (2 μg/ml). Anti-CD3/CD28 dynabeads served as positive control. IFNγ producing T cells were detected by ELISpot after 24 h incubation. Representative images of ELISpot wells were shown. (B) The number of IFNγ-producing T cells was quantified as the spot-forming units (s.f.u.) per 10^6^ PBMCs. (C-G) PBMCs were stimulated with spike peptide pool (2 μg/ml) for 12 h. Activation-induced markers (AIM) CD69 and 4-1BB were measured by flow cytometry. (C) Representative FACS plots showing the SARS-CoV-2-specific AIM^+^ CD8^+^ T cells. (D) Percentages of AIM^+^ CD8^+^ T cells were summarized. (E) T_N_ (CD45RA^+^CCR7^+^), T_EM_ (CD45RA^−^CCR7^−^), T_CM_ (CD45RA^−^CCR7^+^), T_ERMA_ (CD45RA^+^CCR7^−^) populations in AIM^+^ CD8^+^ T cells. (F, G) SARS-CoV-2-specific CD3^+^CD4^−^CD8^−^ (DN) T cells were identified by CD69 and 4-1BB after spike peptide pool stimulation. (H, I) Intracellular cytokine staining assays were performed to confirm the presence of SARS-CoV-2-specific CD8^+^ T cells in PBMCs by measuring intracellular IFNγ expression after peptide pool stimulation. Comparisons were done by Student’s paired-T test in panels B, D and G and one-way ANOVA corrected for multiple comparison in panel I. ****p<0.0001; ns, not significant.

Next, SARS-CoV-2-specific CD8^+^ were identified by AIM T cell assay, based on the previous observation that antigen-specific CD8^+^ T cells could be identified by CD69^+^4-1BB^+^ after the peptide pool stimulation^26^. In line with ELISpot, CD8^+^CD69^+^4-1BB^+^ T cells were seen on d180, and further elevated by 2.7 folds following a booster shot (Fig. 3C and D). Subsequent analysis on phenotypic markers revealed that the majority of the SARS-CoV-2-specific CD8^+^ T cells were CD45RA^+^CCR7^−^ terminally differentiated effector memory cells (T_EMRA_), whereas CD45RA^−^CCR7^−^ effector memory T cells (T_EM_) accounted for only one-fourth of the total antigen-specific CD8^+^ T cells (Fig. 3E), similar to that from recovered COVID-19 patients^26^. The subsets of total CD8^+^ T cells were not affected (Supplementary Fig. 4 and Fig. 5). The presence of antigen-specific CD8^+^ T cells was further confirmed by conventional intracellular cytokine staining (ICS) assay, in which PBMCs were stimulated with spike peptide pool and IFNγ expression in CD8^+^ T cells was measured by flow cytometry. CD8^+^ IFNγ^+^ T cells were detected on d56, and decreased over time till the third vaccination, which elevated IFNγ-expressing CD8^+^ T cells to a greater extent when compared to that on d56 (Fig. 3H and I).

SARS-CoV-2-specific CD4^+^ T cells were identified by activation markers OX40 and 4-1BB after the peptide stimulation^26^. CD4^+^OX40^+^4-1BB^+^ T cells were detected at a low level on d180. The third dose induced the expansion of these CD4^+^ T cells by 5.9 folds within 1 week (Fig. 4A and B). Phenotypic analysis showed that the majority of the SARS-CoV-2-specific CD4^+^ T cells were CD45RA^−^CCR7^+^ T_CM_ and CD45RA^−^CCR7^−^ T_EM_, different from that of SARS-CoV-2-specific CD8^+^ T cells (Fig. 4C). Among SARS-CoV-2-specific CD4^+^ T cells, 57.7% of the cells were CXCR5^+^ cTFH cells (Fig. 4D-G). Among total CD4^+^ T cells, we observed the slightly decreased T_N_ cells, with increased T_EM_ and T_CM_ cells in the peripheral blood (Supplementary Fig. 4 and Fig. 5). Again, SARS-CoV-2-specific CD4^+^ T cells were confirmed by ICS assay. CD4^+^ IFNγ^+^ T cells showed a similar trend as that of CD8^+^ IFNγ^+^ T cells. As expected, the drop of antigen-specific CD4^+^ T cells was reversed by the third dose (Fig. 4H and I).

**Figure 4.**
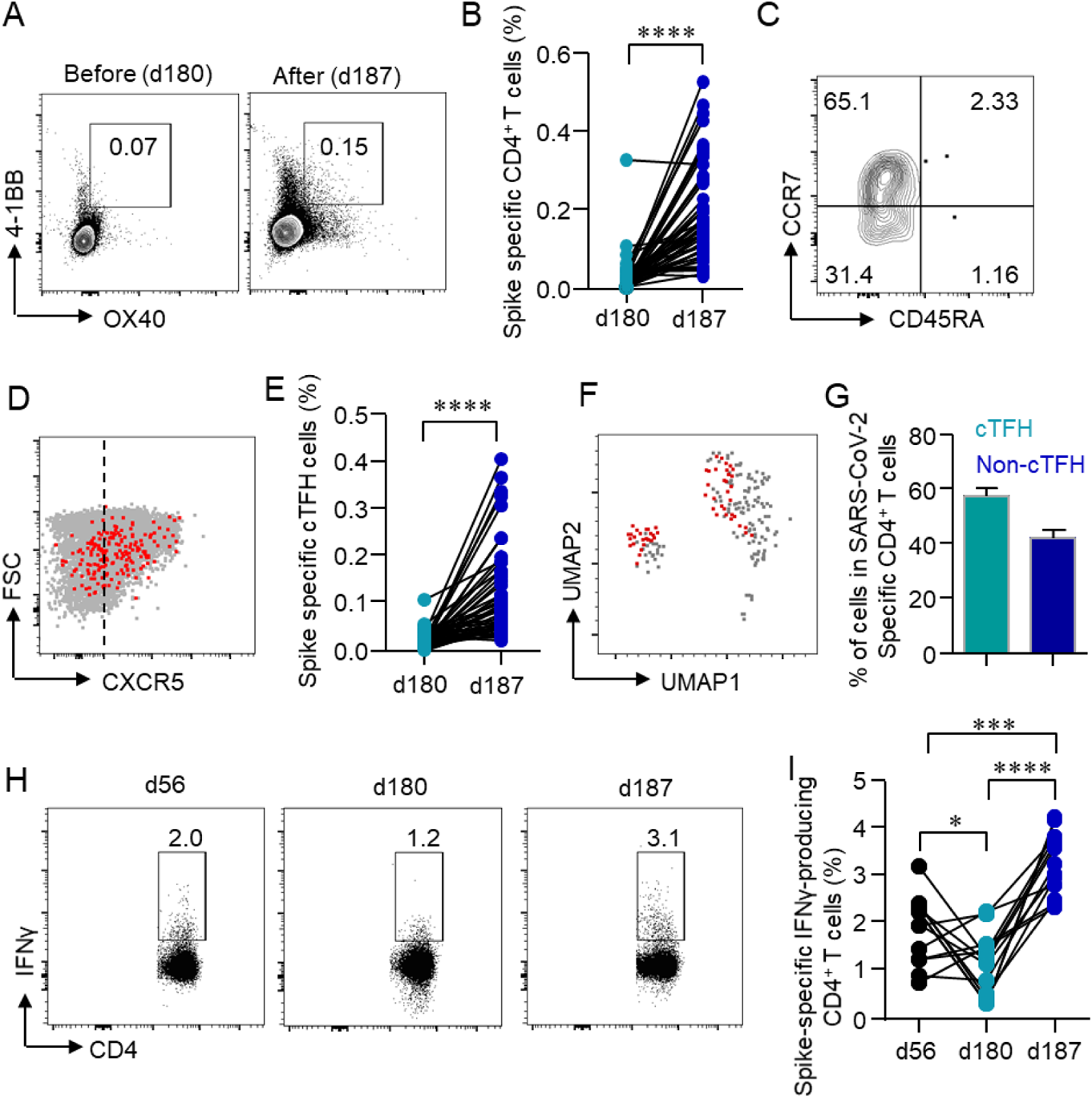
Rapid induction of CD4^+^ T cells by the third dose. (A, B) Activation-induced markers (AIM) OX40 and 4-1BB in CD4^+^ T cells were detected after incubating PBMCs with spike peptide pool (2 μg/ml) for 12 h. Percentages of AIM^+^ CD4^+^ T cells in the PBMCs were summarized. (C) T_N_ (CD45RA^+^CCR7^+^), T_EM_ (CD45RA^−^CCR7^−^), T_CM_ (CD45RA^−^CCR7^+^), T_ERMA_ (CD45RA^+^CCR7^−^) populations in AIM^+^ CD4^+^ T cells were gated. (D) Expression of CXCR5 in AIM^+^ CD4^+^ T cells was measured by flow cytometry. (E) Percentages of circulating follicular helper T cells (cTFH) in total CD4^+^ T cells were summarized. (F) UMAP plot showed the distribution of cTFH cells among AIM^+^ CD4^+^ T cells. (G) Percentage of cTFH cells in AIM^+^ CD4^+^ T cells. (H, I) Intracellular cytokine staining assays were performed to confirm the presence of SARS-CoV-2-specific CD4^+^ T cells in PBMCs by measuring intracellular IFNγ expression after peptide pool stimulation. Comparisons were done by Student’s paired-T test in panels B and E and one-way ANOVA corrected for multiple comparison in panel I. *p<0.05, ***p<0.001, ****p<0.0001.

Besides CD4^+^ and CD8^+^ T cells, we also found a unique population, SARS-CoV-2-specific CD3^+^CD4^−^CD8^−^ T cells persisted 5 months after the second immunization with inactivated vaccines, which were further elevated by the third vaccination (Fig. 3F and G).

### A third dose of inactivated SARS-CoV-2 vaccine boosts immune responses in individuals with negative serologic responses to 2 doses

The weakened or negative serological response has been noted in a small group of vaccinees who had no known immunodeficiency disease or were not in an immune-compromised condition. It is unclear that whether these people are true “non-responders”, and whether a third dose is necessary and beneficial as in immune-compromised patients^33, 34^. In this study, we found NAbs were below the threshold in 9 participants after the primary 2-dose regimen. Of note, these participants have no known immunodeficiency disease or are not in an immune-compromised condition. The level of NAbs in all of the 9 participants increased rapidly within 1 week and reach the peak 2 weeks after the booster shot (Fig. 5A), though NAbs levels in these 9 participants were still significantly lower than others who were seropositive after the second dose (Fig. 5B).

**Figure 5.**
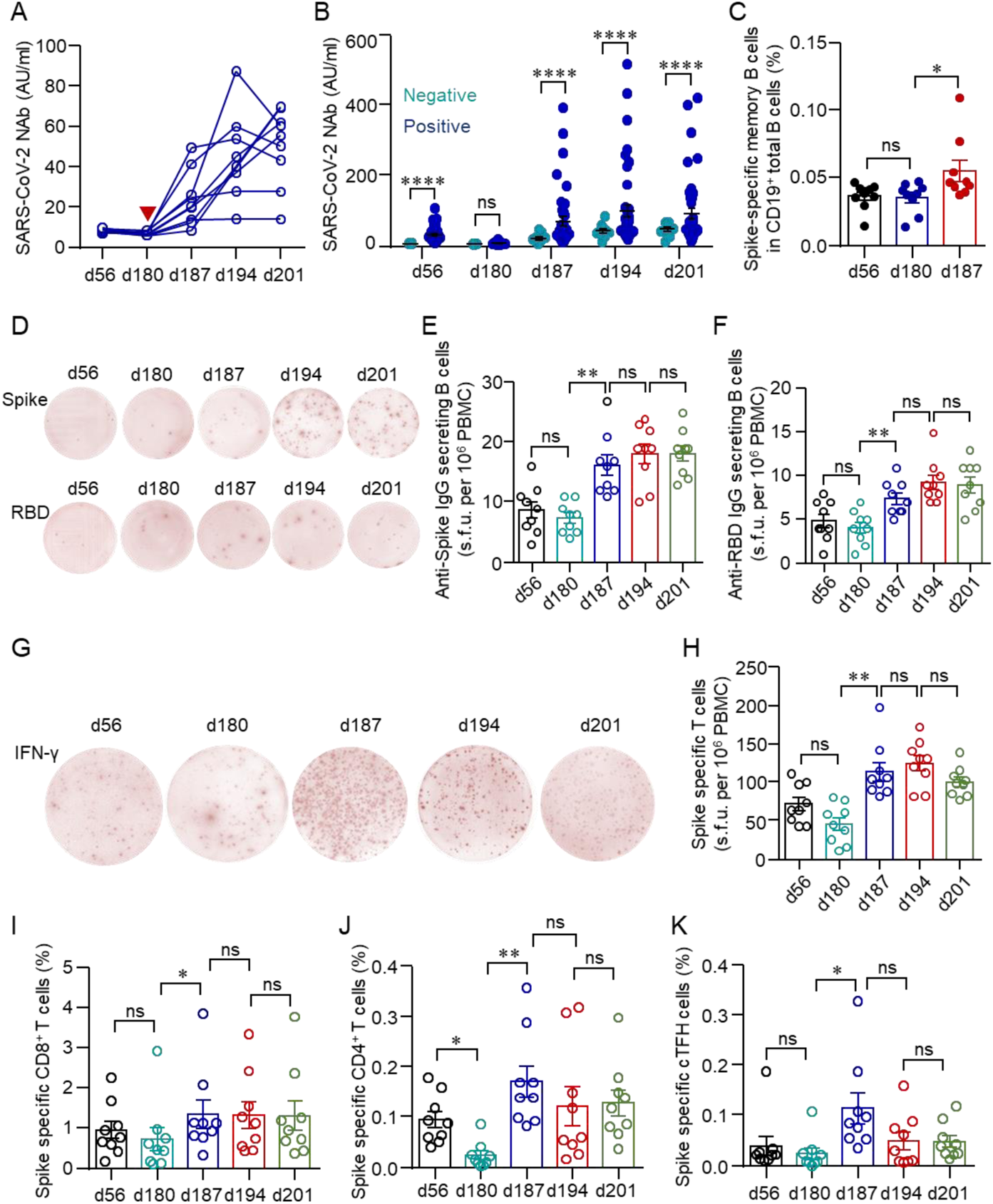
Immune responses in healthcare workers with minimal serologic response to a standard two-dose vaccination schedule. (A) Serum NAbs in HCWs with minimal serologic response to a standard two-dose vaccination schedule were analyzed longitudinally at different time points before and after the third dose (red arrow). (B) Comparison of NAbs between vaccinees with negative or positive serological responses to a standard two-dose vaccination schedule. (C) Percentages of spike-specific memory B cells in HCWs with a negative serological response to a standard two-dose vaccination schedule. (D-F) Numbers of spike- or RBD-specific IgG-producing B cells in HCWs with a negative serological response to a standard two-dose vaccination schedule. (G, H) Numbers of spike-specific IFNγ producing T cells as detected by ELISpot in HCWs with a negative serological response to a standard two-dose vaccination schedule. (I-K) Spike-specific T cells, including CD69^+^4-1BB^+^CD8^+^ T cells, OX40^+^4-1BB^+^CD4^+^ T cells and CXCR5^+^OX40^+^4-1BB^+^ cTFH in HCWs with a negative serological response to a standard two-dose vaccination schedule. Comparisons were done by Wilcoxon rank sum test in B and One-way ANOVA corrected for multiple comparison in the rest panels. *p<0.05, **p<0.01, ****p<0.0001; ns, not significant.

The rapid recall of humoral responses indicated that the immune memory had been successfully established in these seronegative people after the second dose and persisted for at least 5 months. As expected, SARS-CoV-2-specific memory B cells were indeed detected by labeling antigen-binding BCR in these individuals on d56 and d180. The percentage of SARS-CoV-2-specific memory B cells was quite stable over time between d56 and d180, and increased significantly after the third dose (Fig. 5C). ELISpot was next enrolled to identify functional memory B and T cells. Although no NAbs were detected at any time point before the third dose, spike- or RBD-specific IgG-secreting memory B cells were able to be detected by ELISpot on d56 and maintained at a similar level till d180. The number increased within 1 week after the third dose and persisted thereafter (Fig. 5D and F).

Spike-specific T cells were also able to be detected on d56 and d180 as measured by ELISpot, albeit with a downward trend (Fig. 5G and H). As expected, the third dose was capable of enhancing T cell responses in this population (Fig. 5H). The enhancement on SARS-CoV-2-specific CD4^+^ and CD8^+^ T cells by the third dose was further confirmed by measuring AIM^+^ T cells (OX40^+^4-1BB^+^ for CD4^+^ T cells and CD69^+^4-1BB^+^ for CD8^+^ T cells) by flow cytometry. Both spike-specific CD4^+^ T cells and CD8^+^ T cells were successfully expanded (Fig. 5I and J). We also observed the expansion of SARS-CoV-2-specific cTFH cells in these participants (Fig. 5K). Taken together, these results suggested that these participants were not true “no responders”, immune memory had been established but repeated antigen stimulations were needed.

## Discussion

In this study, we investigated the duration of B cell and T cell immunity following the primary 2-dose vaccination regimen of the inactivated SARS-CoV-2 vaccine BBIBP-CorV and the potential benefit of a third dose in a non-randomized trial. Data in this study showed that neutralizing antibodies gradually decreased after the second dose during a 5-month observation. SARS-CoV-2-specific B cells and T cells were detected and persisted in the peripheral blood 5 months after the second vaccination. Both humoral and cellular responses were rapidly and robustly elevated by a third dose of inactivated SARS-CoV-2 vaccine. The induction of both SARS-CoV-2-specific memory B cells and T cells by the inactivated vaccine supports durable protection, as has been shown in mRNA vaccines and other settings^35, 36^. Another key observation in this study was the successful induction of antigen-specific memory B cell and T cell response in HCWs with low serological response to 2 doses of inactivation vaccines.

The NAb titers have been noted to decay following vaccination or SARS-CoV-2 infection^37–39^. However, HCWs who have recovered from COVID-19 show a substantially lower risk of reinfection with SARS-CoV-2^40^. The number of RBD-specific memory B cells remained relatively stable between 6 and 12 months after SARS-CoV-2 infection and BCR repertoire expanded markedly in these recovered patients after receiving an mRNA vaccine^41^. Consistently, we found that spike- and RBD-specific memory B cells could still be detected in all COVID-19 convalescent patients up to 13 months after infection (Supplementary Fig. 6). SARS-CoV-2 mRNA vaccines were also capable of inducing sustained immune memory and long-term protecting effects^35, 42, 43^. Our study, together with these aforementioned studies, suggests that SARS-CoV-2-specific memory B cells may sustain longer than plasma cells. Prediction of vaccine efficacy should not solely rely on NAbs titers. Rather, memory B cells should be taken into account whenever it is possible.

Recently, a third dose of inactivated vaccine CoronaVac elicited a rapid and long-lasting recall antibody response with the capability of neutralizing several VOCs^44^. The rapid and strong secondary response by the inactivated vaccine represents a characteristic secondary immune response, which demonstrates the establishment of long-term immune memory within these participants. Our data confirm that SARS-CoV-2-specific memory B cells persisted 6 months later the primary vaccination and expanded substantially by a third dose of vaccine. The secondary immune response is one of the most important features of immune memory stimulated by vaccination, which is characterized by a faster and stronger immune response by memory B cells or plasma cells^45^. Despite the waning of antibodies, the generation of SARS-CoV-2-specific memory B cells could mediate recall response in future infection and provide protective immunity. As has been observed in recovered COVID-19 patients, long-lived bone marrow spike-binding plasma cells are quiescent^27^, which explains the declining antibodies over time. Nonetheless, further investigation should be conducted on how waning immunity could affect protection against COVID-19.

Our data revealed that long-lasting memory B cells formed a basis for an enormous lift of serum NAbs after the third dose, not only against WT SARS-CoV-2 but also against B.1.617.2 Delta variant. As shown in Supplementary Fig.1, the inactivated vaccine induced a broad spectrum of antibody responses to multiple structural proteins of the virus. Whether the lifted cross-reactive NAbs still targeted to RBD, or other domains of S protein, or even other proteins were involved, is an interesting question meriting further study.

Our study showed that SARS-CoV-2-specific CD4^+^ and CD8^+^ T cells in convalescent COVID-19 patients could persist up to 13 months (Supplementary Fig. 7). SARS-CoV-2-specific CD4^+^ and CD8^+^ T cells induced by BBIBP-CorV were detected in all the HCWs who received 2 doses of vaccine and last for at least 5 months. In line with previous data from an mRNA vaccine^20^, SARS-CoV-2-specific CD4^+^ T cells induced by inactivated vaccine mainly fall into central and effector memory subsets, while the majority of SARS-CoV-2-specific CD8^+^ T cells were T_EMRA_ cells. Because of the high CCR7 expression, the central memory T cells reside in the secondary lymphoid organs and readily expand upon infection or booster vaccination. Effector memory T cells circulate in between blood and peripheral tissues, providing prompt immune response at the early time point of infection^46, 47^. For the emergence of VOCs of SARS-CoV-2, there is growing concern over vaccine efficacy. In addition, T cells from COVID-19 convalescents and in recipients of mRNA vaccine could recognize epitopes throughout the SARS-CoV-2 spike protein and the total reactivity against SARS-CoV-2 variants of B.1.1.7, B.1.351, P.1, and CAL.20C lineages were mostly maintained in terms of magnitude and frequency of response^48^. SARS-CoV-2-specific T cells from convalescent COVID-19 patients or vaccinated individuals targeted the conserved epitopes between prototype and VOCs^49, 50^. Thus, SARS-CoV-2-specific T cells are less likely to be affected by antibody escapes mutations in VOCs. Moreover, the inactivated virus vaccine comprises all viral structural proteins. This means more epitopes, especially those conserved epitopes in proteins other than spike are engaged as compared to mRNA or recombinant protein vaccines involving only RBD or spike. Ideally, a potent adjuvant rather than aluminum salts should be used to fully realize this advantage.

Another interesting finding of this study was the induction of SARS-CoV-2-specific cTFH cells by the inactivated vaccine. Our previous study has demonstrated the mobilization of cTFH cells after the first dose of inactivated vaccination^11^. For the relatively late development of humoral response by the inactivated vaccines, early response of cTFH cells could contribute to the early protection of vaccination when antibody level was still low^3, 51^. cTFH cells have been associated with humoral response in COVID-19 patients or vaccination individuals^20, 23^. Data in this study further confirmed that the induction of cTFH cells by the inactivated vaccine correlated with humoral response closely.

A third dose of the SARS-CoV-2 vaccine may be particularly in need for the immunocompromised population. Low seroconversion rates to mRNA vaccination have been reported in solid transplant recipients^52^. The applications of immune suppressants or B cell-targeted therapies could also hamper serological response to vaccination^53, 54^. Recently, data show that antibody response was enhanced in kidney transplant recipients with minimal serologic response to 2 doses by a third dose of mRNA vaccine^33^. In this study, we found that a group of healthy individuals was not mounted with detectable NAbs by two doses of vaccination. However, memory B and T cells could be detected 5 months after the second immunization. The third dose of inactivated vaccine could induce an elevated level of NAbs and an extensive expansion of memory cells within 1 week, suggesting that the memory response could also be effectively recalled upon SARS CoV-2 infection. Therefore, these individuals were not true “non-responders” but already equipped with immune memory which may at least confer protection against severe diseases. Nevertheless, it should be noted that further studies are needed to confirm the relevance between these memory responses and protection. Moreover, a third dose is necessary to further secure the protection.

In conclusion, our study revealed sustained memory B and T cell responses after a standard 2-dose vaccination regimen of inactivated vaccines. Immune responses against SARS-CoV-2 were readily recalled and further elevated by a third dose of the inactivated vaccine, which is especially beneficial for individuals facing high exposure risks, such as HCWs, and has been demonstrated to be safe and effective in our study and others^33, 55^.

## Author Contributions

HX, HZ and JW supervised the study. YL, QZeng, CD, MLi, LL and DL contributed equally to the study. HX, HZ, JW and YL conceived and designed the study. YL, DL, JM and RM recruited participants in the trial and collected blood samples. QZeng, CD, ML, MLi and LL performed the experiments and collected data. QZeng and QZhou performed statistical analysis. HZ, YL, QZeng, CD, MLi, LL and DL drafted the report. HX, JW, SP and MLiu made critical revision on the manuscript. All authors contributed to the interpretation of data. All authors approved the final version before submission.

## Data Availability

All data were included in the current form of manuscript.

## Acknowledgment

We thank Prof. Lu Lu at Fudan University for his kind help in pseudovirus assays. The work is supported by The Talent Program of the First Affiliated Hospital, Sun Yat-sen University (Y70311) and The Hundred Talent Program of Sun Yat-sen Univetsity (Y61229).

