## Supplementary Material for "Robust induction of B cell and T cell responses by a third dose of inactivated SARS-CoV-2 vaccine"

**Supplementary Table 1 Demographics and vaccination-related adverse events**

| | | Morning group | Afternoon group | Total |
|---|---|---|---|---|
| Participants, N | | 25 | 25 | 50 |
| Age, mean (SD) | | 26.88 (4.86) | 27.88 (6.93) | 27.38 (5.94) |
| Gender, male (%) | | 9 (36.0) | 11 (44.0) | 26 (41.3) |
| Adverse events, | N (%) | 5 (20.0%) | 1 (4.0%) | 6 (12.0%) |
| Injection site symptoms | Induration, N (%) | 1 (4.0%) | 0 (0.0%) | 1 (2.0%) |
| | Swollen, N (%) | 1 (4.0%) | 0 (0.0%) | 1 (2.0%) |
| Systematic symptoms | Fever, N (%) | 2 (8.0%) | 0 (0.0%) | 2 (4.0%) |
| | Fatigue, N (%) | 2 (8.0%) | 1 (4.0%) | 3 (6.0%) |
| Respiratory symptoms | Runny nose, N (%) | 1 (4.0%) | 0 (0.0%) | 1 (2.0%) |
| | Cough, N (%) | 1 (4.0%) | 0 (0.0%) | 1 (2.0%) |
| | Sore throat, N (%) | 1 (4.0%) | 0 (0.0%) | 1 (2.0%) |

**Supplementary Table 2 Demographics of the COVID-19 convalescent patients**

| Patient ID | Age range | Gender | Days post-diagnosis | NAbs* (AU/ml) |
|---|---|---|---|---|
| #1 | 21-25 | F | 407 | 28.67 |
| #2 | 36-40 | F | 406 | 582.92 |
| #3 | 31-35 | F | 411 | 14.79 |
| #4 | 36-40 | M | 409 | 74.09 |
| #5 | 46-50 | F | 403 | 23.55 |

*NAbs: neutralizing antibodies

**Supplementary Table 3 Antibodies used in this study.**

| Marker | Fluorophore | Vendor | Cat# | Clone |
|---|---|---|---|---|
| CD4 | BV510 | Biolegend | 357420 | A161A1 |
| CD8 | Percp | Biolegend | 344708 | SK1 |
| CD45RA | BV650 | Biolegend | 304136 | HI100 |
| CD3 | Pacific Blue™ | Biolegend | 300330 | HIT3a |
| CD137 | PE | Biolegend | 309804 | 4B4-1 |
| OX40 | BV605 | Biolegend | 350028 | ACT35 |
| CD69 | Super Bright 436 | Invitrogen | 62-0699-42 | FN50 |
| CCR7 | APC/cy7 | Biolegend | 353212 | G043H7 |
| CD19 | PE/cy7 | Biolegend | 302216 | HIB19 |
| PD-1 | PE/cy7 | Biolegend | 367414 | NAT105 |
| CXCR5 | BV711 | Biolegend | 356934 | J252D4 |
| CD27 | AF700 | Biolegend | 356416 | M-T271 |
| IgD | FITC | Biolegend | 348206 | IA6-2 |
| CD38 | BV650 | Biolegend | 356620 | HB-7 |
| IgM | BV605 | Biolegend | 314524 | MHM-88 |
| IgG | AF647 | Biolegend | 302216 | HIB19 |
| IgA | PE | Biolegend | 333504 | HM47 |
| IFNγ | PE-CY7 | Biolegend | 502528 | 4S.B3 |
| Zombie | Zombie Red | Biolegend | 423110 | —— |
| Streptavidin | BV421 | Biolegend | 405226 | —— |
| Streptavidin | BV510 | Biolegend | 405233 | —— |
| Streptavidin | BV785 | Biolegend | 405249 | —— |

**Supplementary Figures and legends**

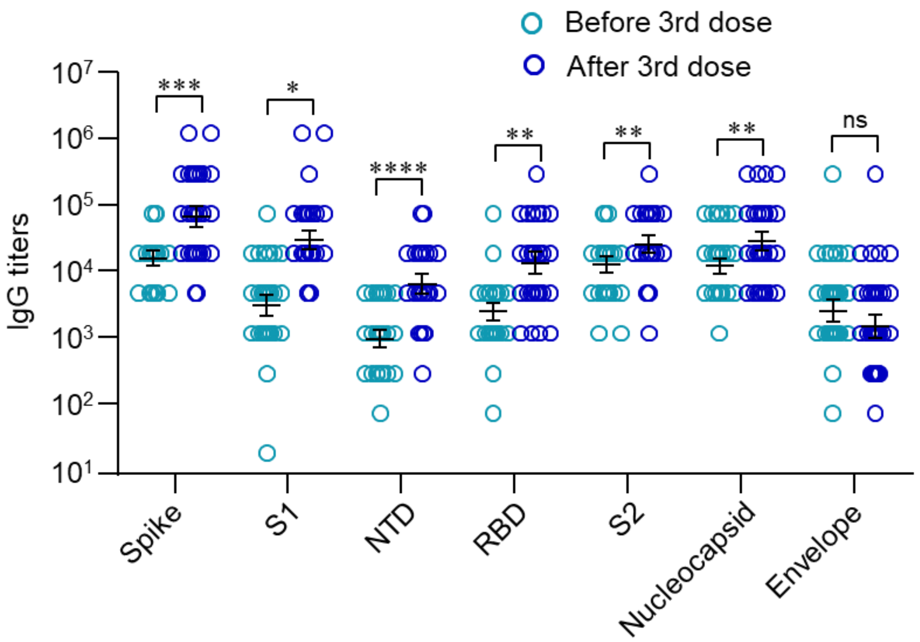


**Supplementary Fig. 1 Antibody spectrum induced by inactivated vaccine BBIBP-CorV**. Study was carried out as in Fig.1. IgG against SARS-CoV-2 spike, spike S1, spike N terminal domain (NTD), receptor-binding domain (RBD), spike S2, nucleocapsid and envelope protein before (d180) and after (d194) the third dose were measured by ELISA. ns, not significant, *p<0.05, **p<0.01, ***p<0.001 and ****p<0.0001 by paired Student's t test.

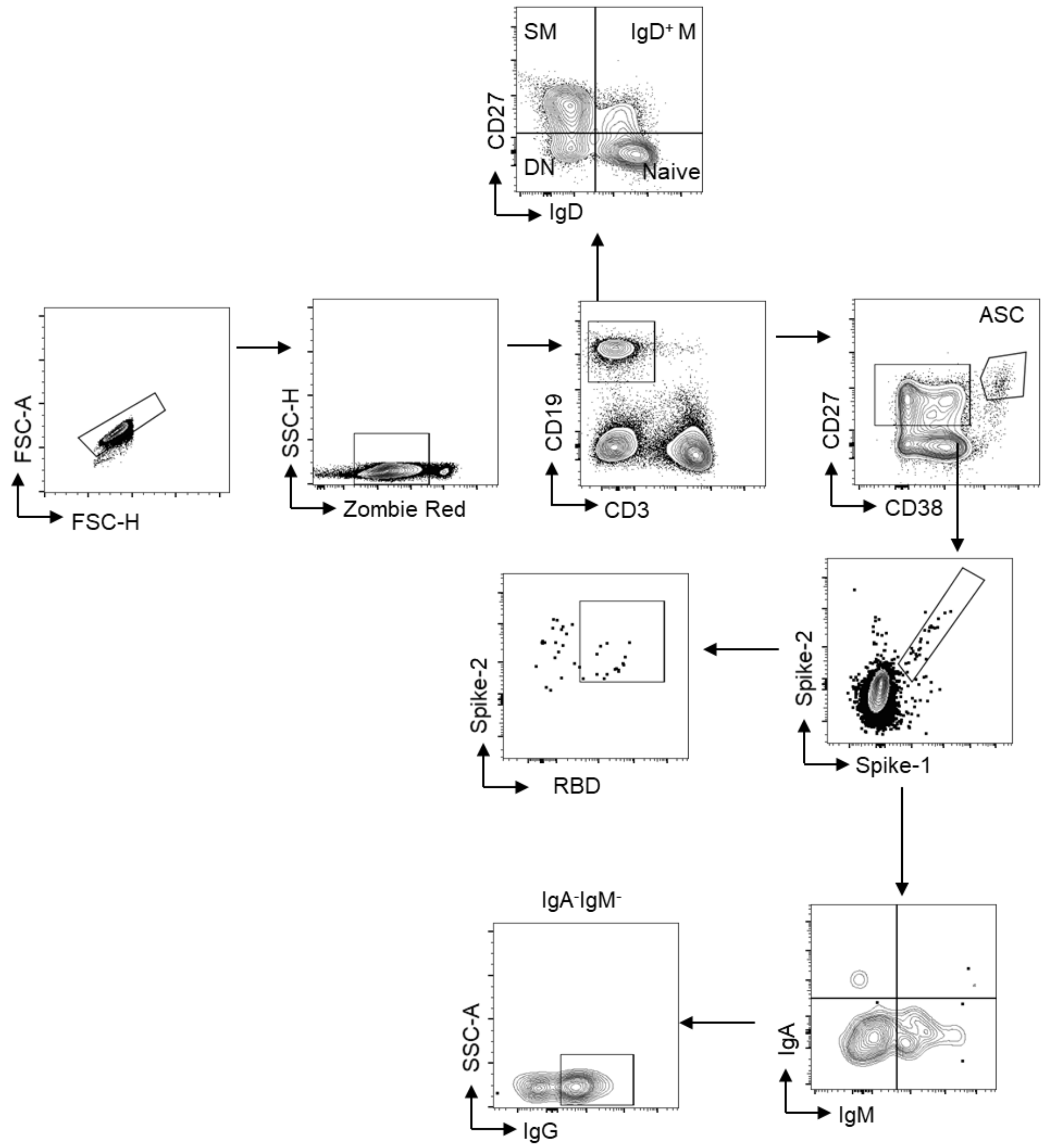


**Supplementary Fig. 2 Gating strategy for B cell analysis**. PBMCs were collected on day (d) 180 before the third dose and d187 after the third dose of vaccination. Cells were stained with antibodies against CD3, CD19, CD27, IgD, IgM, IgG, IgA and CD38. Dead cells were first gated out by gating on Zombie Red$^-$ population. Cells were then gated on CD19$^+$ B cells for further analysis: naïve B cells: IgD$^+$CD27$^-$, IgD$^+$ memory B cells: IgD$^+$CD27$^+$, switched memory B cells: IgD$^-$CD27$^+$, double-negative (DN) B cells: IgD$^-$CD27$^-$, antibody-secreting cells (ASC): CD38$^+$CD27$^+$.

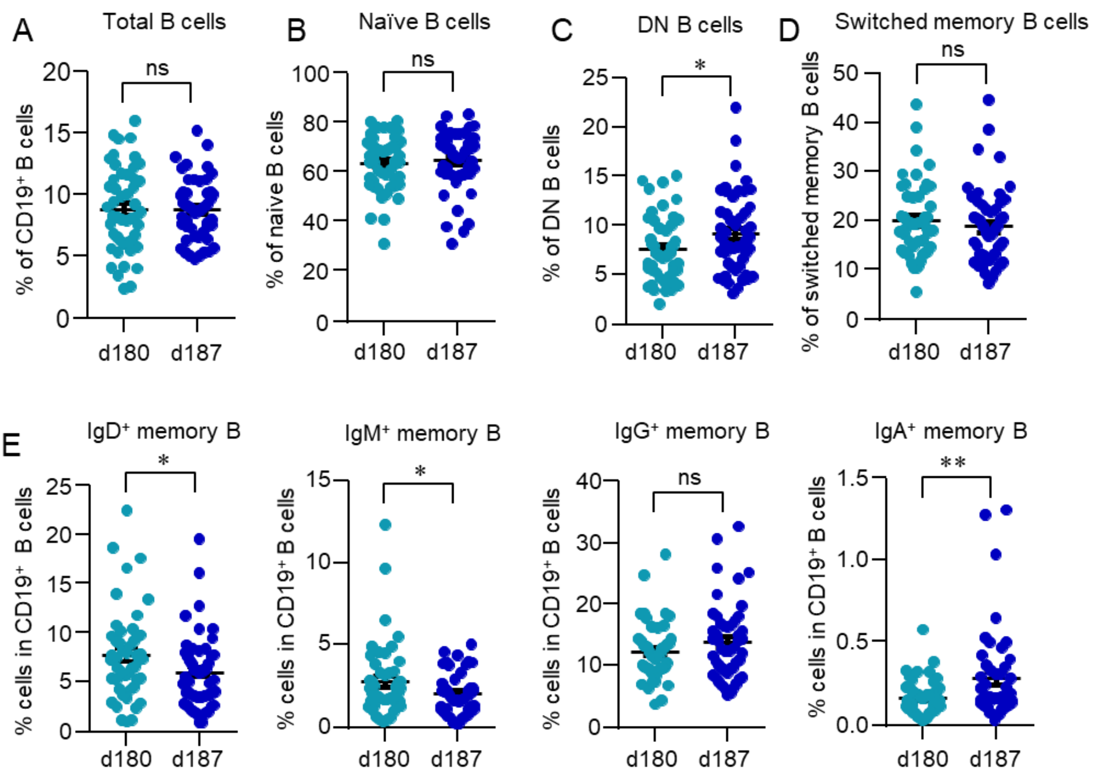


**Supplementary Fig. 3 B cell response to a third dose of inactivated SARS-CoV-2 vaccine**. Blood samples were collected on day (d) 180 and d187. Data were acquired by flow cytometry. The gating strategy is shown in Supplementary Fig. 2. B cell subsets were summarized and shown as dot plots. Data are expressed meat ± SEM. ns, not significant, *p<0.05, **p<0.01. Comparisons were done by Student's t test.

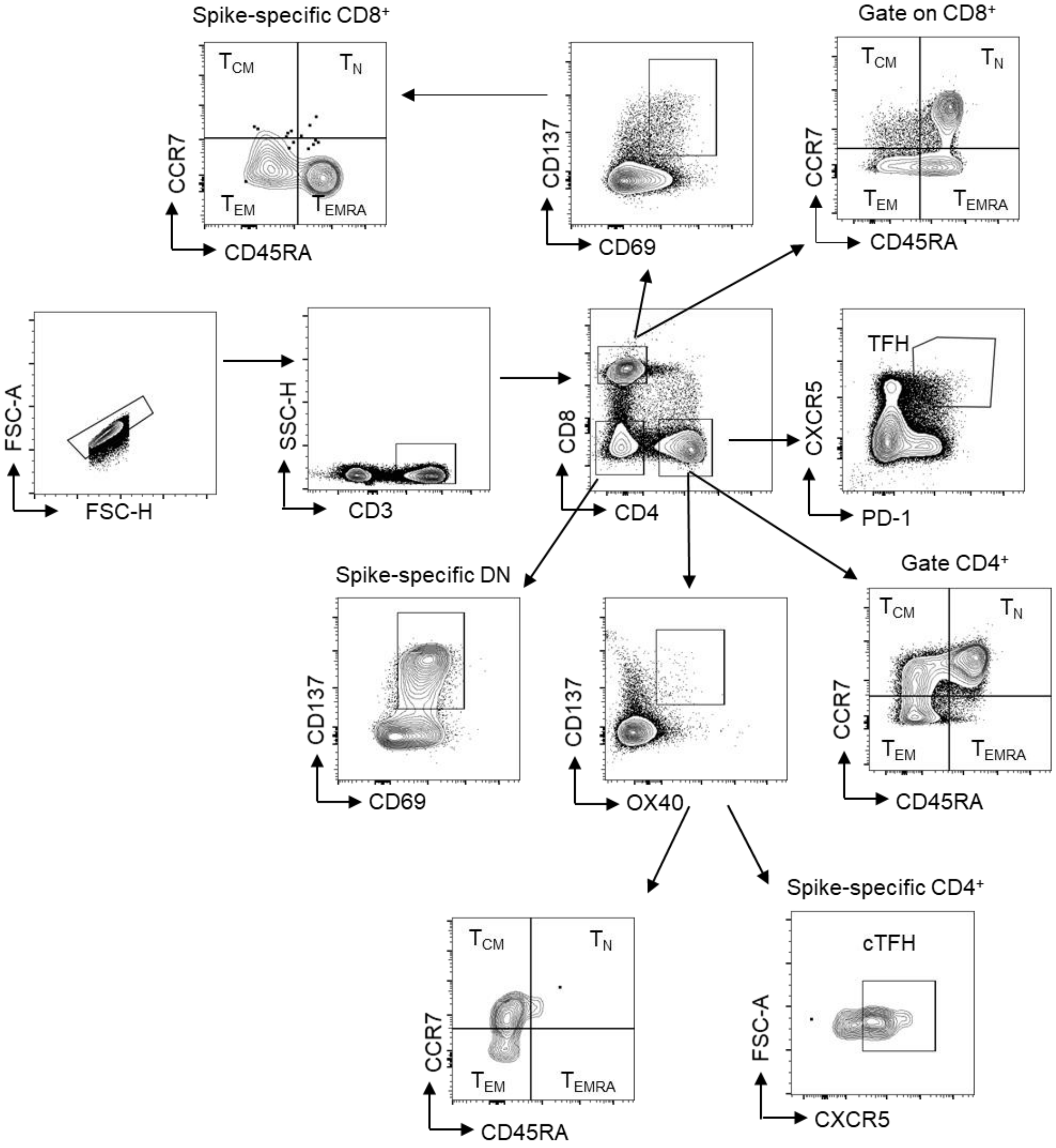


**Supplementary Fig. 4 Gating strategy for T cell analysis**. PBMCs were collected on day (d) 180 before the third dose and d187 after the third dose vaccination. Cells were stained with antibodies against CD3, CD4, CD8, IgD, IgM, IgG, IgA and CD38. T cell phenotyping: TFH cells: $CD4^{+}CXCR5^{+}PD\text{-}1^{+}$, Spike-specific $CD4^{+}$ T cells: $OX40^{+}4\text{-}1BB^{+}$, Spike-specific $CD8^{+}$ T cells: $CD69^{+}4\text{-}1BB^{+}$, naïve T cells ($T_N$): $CD45RA^{+}CCR7^{+}$, effector memory T cells ($T_{EM}$): $CD45RA^{-}CCR7^{-}$, central memory T cells ($T_{CM}$): $CD45RA^{-}CCR7^{+}$, terminally differentiated T cells ($T_{EMRA}$): $CD45RA^{+}CCR7^{-}$.

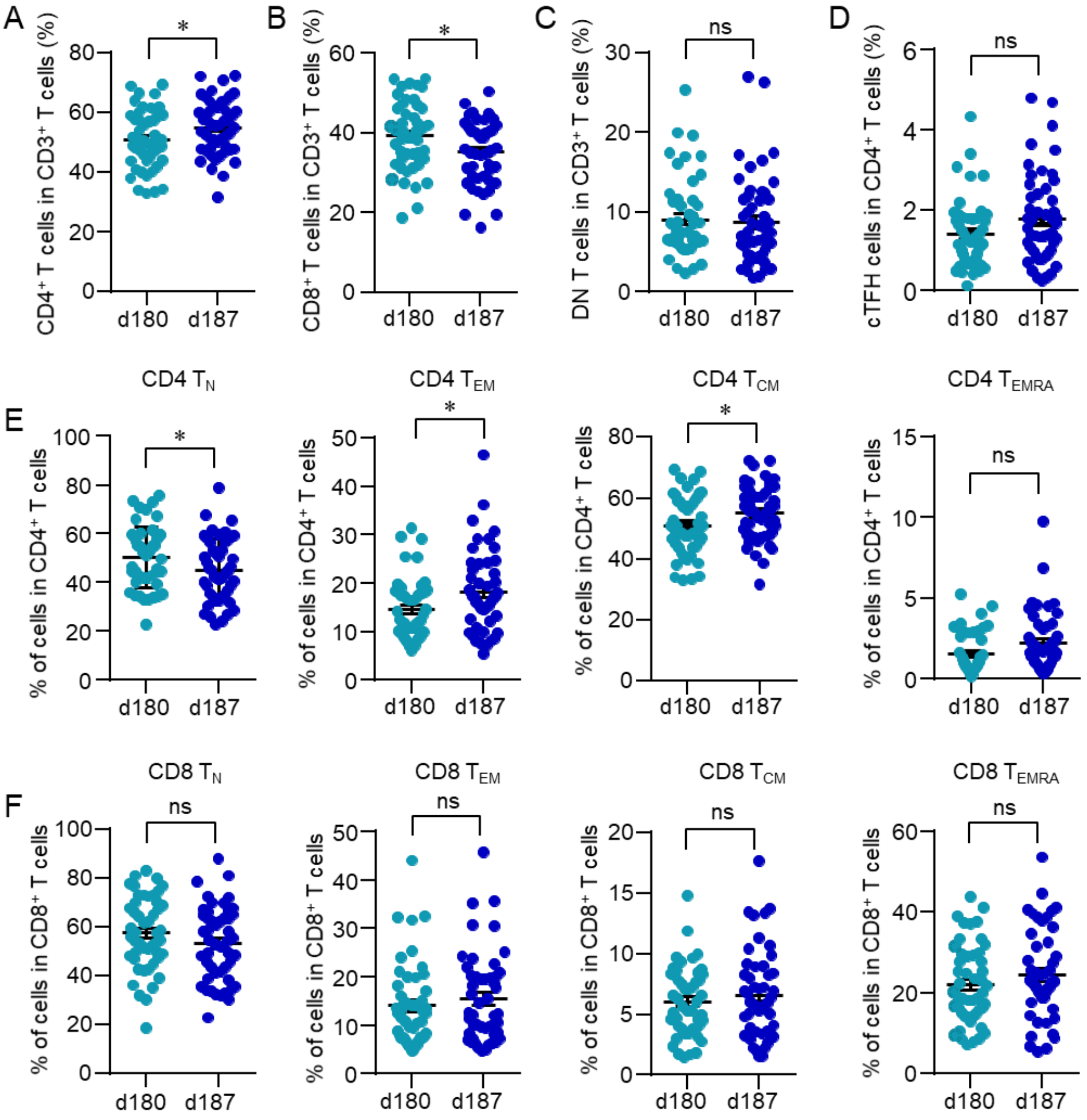


**Supplementary Fig. 5 T cell response to a third dose of inactivated SARS-CoV-2 vaccine**. Blood samples were collected on day (d) 180 and d187. Data were acquired by flow cytometry. The gating strategy is shown in Supplementary Fig. 4. T cell subsets were summarized and shown as dot plots. Data are expressed mean ± SEM. ns, not significant, *$p<0.05$, Comparisons were done by Student's t test.

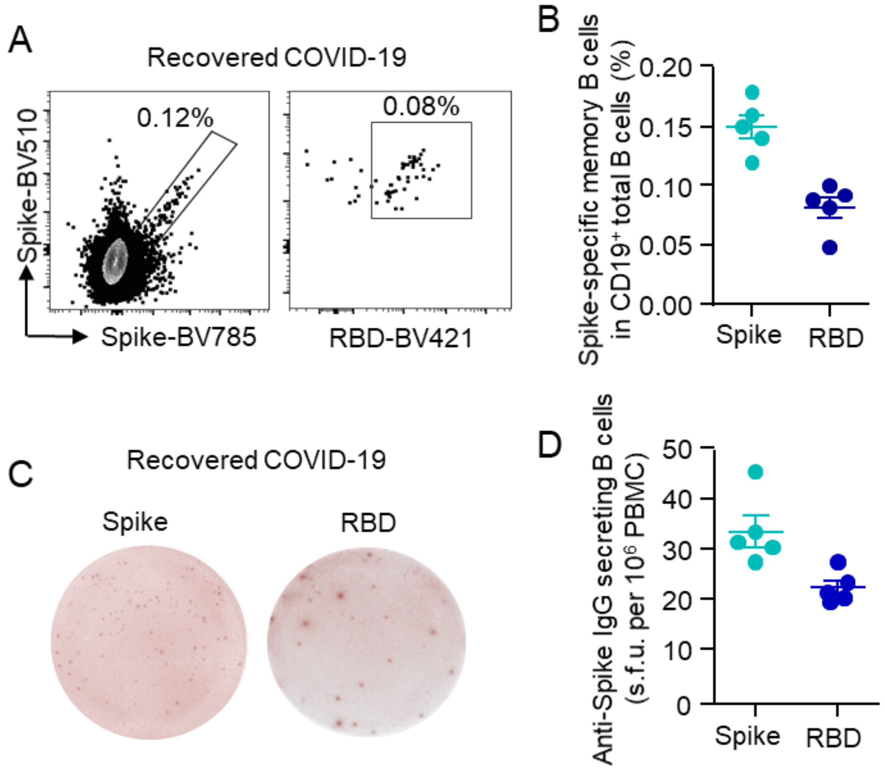


**Supplementary Fig. 6 SARS-CoV-2-specific memory B cells in COVID-19 convalescent patients**. PBMCs were isolated from patients who have recovered from COVID-19 over 13 months. (A, B) Spike- and receptor-binding domain (RBD)-specific memory B cells were measured by flow cytometry. Representative FACS plots showing spike- and RBD-specific memory B cells. Data were summarized from 5 independent samples. (C, D) SARS-CoV-2-specific memory B cells were further detected by ELISpot. Images of ELISpot wells were shown. Spot-forming units (s.f.u) were summarized and adjusted to $10^6$ PBMCs.

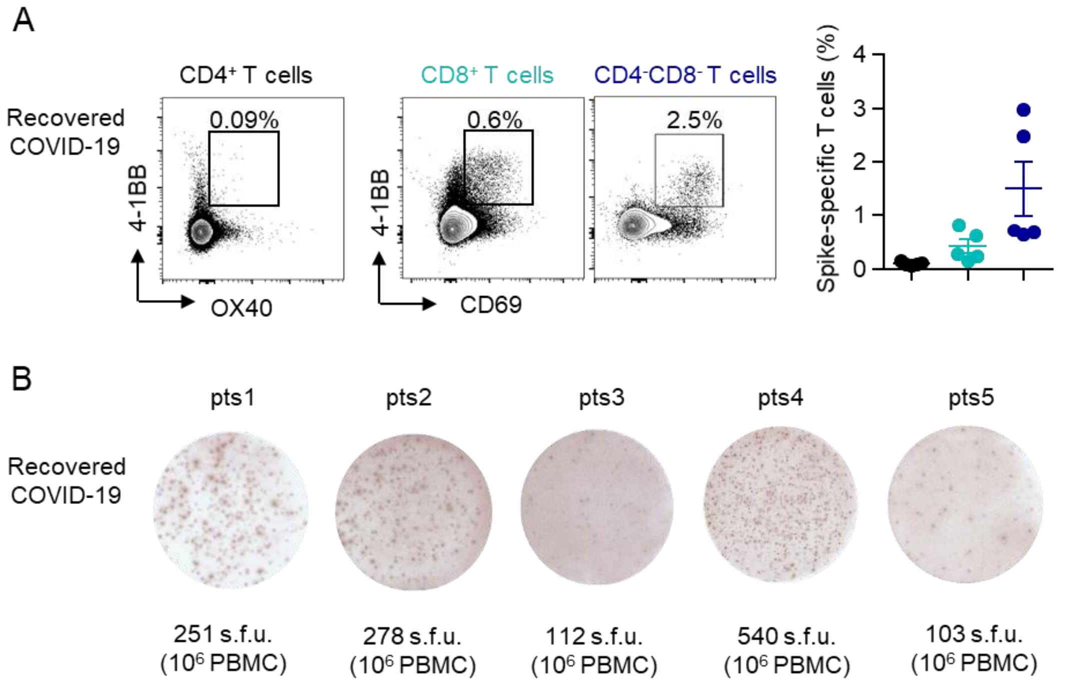


**Supplementary Fig. 7 SARS-CoV-2-specific T cells in convalescent COVID-19 patients**. (A) PBMCs were stimulated with spike peptide pool (2 μg/ml) for 12 h. SARS-CoV-2-specific CD4$^+$ T cells (OX40$^+$4-1BB$^+$), CD8$^+$ T cells (CD69$^+$4-1BB$^+$) and CD3$^+$CD4$^-$CD8$^-$ T cells (CD69$^+$4-1BB$^+$) were measured by flow cytometry. Data were summarized from 5 independent samples. (B) SARS-CoV-2-specific T cells were further detected by IFNγ ELISpot. Images of ELISpot wells of each sample were shown. Spot-forming units (s.f.u) were summarized and adjusted to 10$^6$ PBMCs.
